# Associations Between Prenatal Cannabis Exposure and Birth Outcomes: Results from a Prospective Cohort Study

**DOI:** 10.64898/2026.03.01.26347369

**Authors:** Anna Constantino-Pettit, Cassandra Trammel, Arpana Agrawal, Christopher Smyser, Ebony Carter, Ryan Bogdan, Cynthia Rogers

**Affiliations:** Department of Psychiatry, Washington University in St. Louis; Department of Obstetrics & Gynecology, Division of Maternal-Fetal Medicine, University of Michigan; Department of Neurology, Washington University in St. Louis; Department of Pediatrics, Washington University in St. Louis; Department of Radiology, Washington University in St. Louis; Department of Neurosurgery, Washington University in St. Louis; Department of Obstetrics & Gynecology, Division of Maternal-Fetal Medicine, University of North Carolina at Chapel Hill; Department of Psychological and Brain Sciences, Washington University in St. Louis

**Author notes:** **Please address all correspondence to:** Dr. Anna Constantino-Pettit, 4444 Forest Park Ave, # 06528, St. Louis, MO 63110.

## Abstract

**Objective:** Cannabis use during pregnancy is increasing; associations with neonatal growth may be confounded by nicotine. We evaluated prenatal cannabis exposure (PreCE) and neonatal outcomes in a prospective cohort with biochemical control for nicotine exposure.

**Methods:** In the Cannabis Use During Early Life and Development (CUDDEL) study, pregnant women with a lifetime history of cannabis use were classified as PreCE if they self-reported use or had urine THC-COOH positivity at any trimester (n=297) and as unexposed if they reported no use and tested negative (n=151). Linear regression and modified Poisson models estimated associations with birthweight and small for gestational age (SGA; <10th and <5th percentiles), adjusting for sociodemographic factors, gestational age, maternal age and BMI, and urinary cotinine. Analyses stratified by cannabis use frequency (>weekly vs <monthly) and cotinine status.

**Results:** Participants (N=448; 18-41 years; 85.3% non-Hispanic Black) had lower birthweight with PreCE in adjusted models (Beta=-0.08; padj=0.041). High-frequency PreCE was associated with lower birthweight compared with unexposed pregnancies (Beta=-0.13; padj=0.03), whereas low-frequency PreCE was not. Cotinine-positive PreCE showed the greatest birthweight reduction versus unexposed (Beta=-0.20; padj<0.001). PreCE was also associated with higher likelihood of SGA <5th percentile; risk was highest in PreCE+Nicotine compared with both unexposed and PreCE-Nicotine groups.

**Conclusions:** Prenatal cannabis exposure was associated with reduced birthweight and SGA in this cohort. Nicotine co-exposure intensified these associations, yet effects persisted without cotinine, supporting cannabis as an independent perinatal risk factor and emphasizing the value of cotinine assessment in populations where blunt use or secondhand exposure is common.

## OBJECTIVE

Cannabis use during pregnancy is increasingly common,^1^ despite professional recommendations against use^2^. Prenatal cannabis exposure (PreCE) has been associated with reduced birthweight, although prior studies have inadequately controlled for concurrent nicotine use^3^. We examined PreCE associations with neonatal outcomes in a prospective cohort of pregnant women with pre-pregnancy cannabis use, controlling for concomitant nicotine exposure.

## STUDY DESIGN

The ongoing Cannabis Use During Early Life and Development (CUDDEL) longitudinal study recruited women with a lifetime history of cannabis use who reported cannabis use or whose urine tested positive for the cannabis metabolite THC-COOH (11-nor-9-carboxy-tetrahydrocannabinol) at any trimester visit (PreCE; n=297) and those who reported no use and whose urine was THC-COOH negative (nPreCE; n=151; **eTable 1**). Participants provided informed consent to a research protocol approved by the Washington University Institutional Review Board.

Linear regression and modified Poisson models with and without covariates regressed offspring birthweight, small for gestational age (SGA; i.e., ≤10^th^ and ≤5^th^ percentile), and other neonatal outcomes on PreCE (**eTable 2**). Covariates included Area Deprivation Index, infant gestational age at birth, maternal age, body mass index, and presence (yes/no) of urinary cotinine, a marker for nicotine exposure (except when examining PreCE±Nicotine). Follow-up analyses stratified PreCE by nicotine and self-reported frequency of cannabis use (i.e., ≥ weekly use [High PreCE, n=122], ≤ monthly use [Low PreCE, n=166]; **eTable 2**).

## RESULTS

### Demographics

The sample included 448 pregnant women (18-41 years; 382 [85.3%] non-Hispanic Black).

### Birthweight

Low birthweight was associated with PreCE in adjusted models (β= -0.08, *p*_*adj*_=0.041, **Figure 1A**), and varied by frequency of use and nicotine exposure/cotinine presence (**Figure 1B-C, eTable 2**). High frequency PreCE was associated with lower birth weight compared to nPreCE (β= -0.13, *p*_*adj*_=0.03); low frequency PreCE did not significantly differ from nPreCE or High PreCE (**Figure 1B; eTable 2**). PreCE+Nicotine was associated with lower birthweight relative to nPreCe (β= -0.20, *p*_*adj*_<0.001); PreCE-Nicotine did not differ from PreCE+Nicotine or nPreCE (**Figure 1C; eTable 2**).

**Figure 1.**
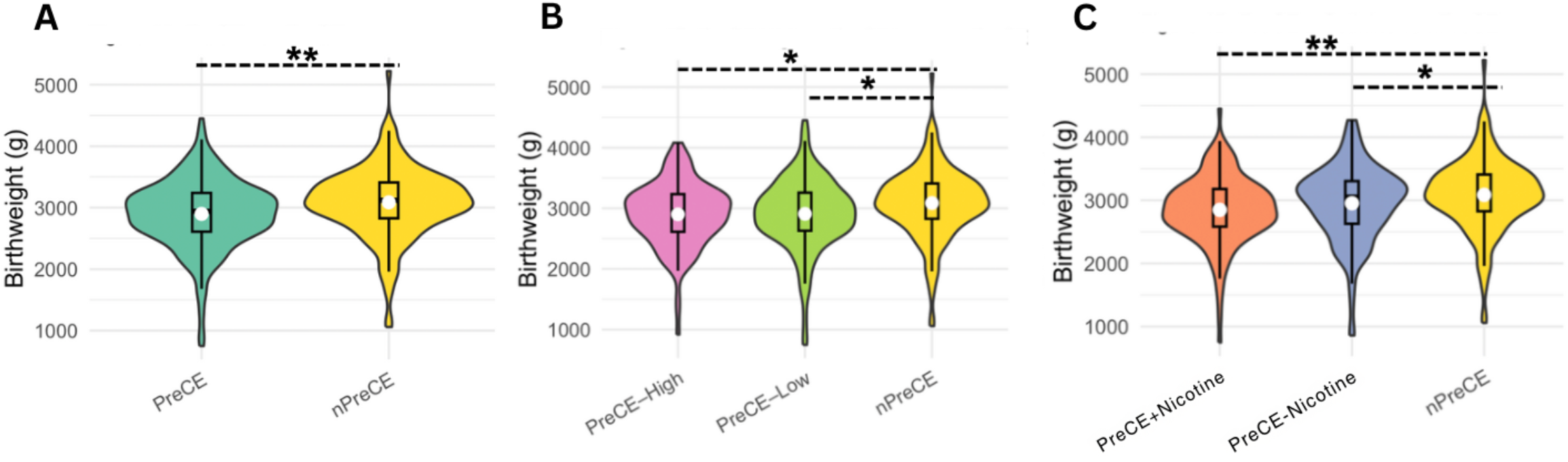
Pairwise Differences in Neonatal Birthweight across Prenatal Cannabis and Nicotine Exposure Groups Note:*p=0.05, **p<0.005. **A**. PreCE (2940 [750-4450]g) vs nPreCE (3120 [1060-5220]g), unadjusted beta = -188.08, SE=57.06, p-value=0.001; adjusted beta = -0.08, SE=0.04, p-value=0.041. **B**. Weekly or greater use (i.e., PreCE-High: 2954 [920-4080]g) vs Monthly or less use (i.e., PreCE-Low: 2940 [750-4450]g), unadjusted beta= -7.52, SE=66.29, p-value=0.910; adjusted beta= -0.02, SE=0.04, p-value=0.650; PreCE-High vs nPreCE: unadjusted beta = -186.01, SE=66.94, p-value=0.006; adjusted beta= -0.13, SE=0.06, p-value=0.025; PreCE-Low vs nPreCE: unadjusted beta = -178.44, SE=65.56, p-value=0.007; adjusted beta= -0.07, SE=0.05, p-value=0.166. **C**. Presence (PreCE+Nicotine: 2860 [750-4450]g) vs Absence (PreCE-Nicotine: 3050 [860-4270]g) cotinine exposure: PreCE+Nicotine vs. PreCE-Nicotine: unadjusted beta = -105.74, SE=67.36, p-value=0.118; adjusted beta=0.07, SE=0.21, p-value=0.732; PreCE+Nicotine vs nPreCE: unadjusted beta = -246.63, SE=66.74, p-value < 0.001; adjusted beta= -0.20, SE=0.05, p-value< 0.001; PreCE-Nicotine vs nPreCE: unadjusted beta= -140.89, SE=72.83, p-value=0.054; adjusted beta= -0.03, SE=0.05, p-value=0.537.

### SGA/LGA

PreCE was associated with a higher likelihood of SGA ≤ 5^th^ percentile (**eTable 2**). Even after adjusments, SGA was more common in PreCE+Nicotine relative to nPreCE and to PreCE-Nicotine.

## CONCLUSION

PreCE is associated with low birthweight and SGA. We extend literature linking PreCE to this neonatal outcome^4^ by providing evidence that these associations may be worsened by, but partly independent of, nicotine exposure. Despite exclusion for self-reported tobacco use, urine testing demonstrated high exposure rates, possibly due to blunt usage (135[45.5%]PreCE) or secondhand exposure (36%) PreCE)) ^5^. While nicotine co-exposure was more common in PreCE-High, it was also noted for PreCE-Low (71[59.8%] vs. 79[48.2%]). This influence of concomitant nicotine and cannabis exposure on birthweight, which may have been obscured without urinary evaluation, highlights the importance of carefully characterizing nicotine co-use in samples where blunt usage is typical, especially as findings suggest that frequency of use and nicotine co-exposure exacerbate birthweight differences. The observed birthweight reductions associated with PreCE – with associations persisting without concurrent nicotine exposure – suggest that cannabis represents a distinct perinatal risk factor requiring dedicated research attention and prenatal counseling.

## Supporting information

Supplemental eTable1, eTable2

## Data Availability

All data produced in the present study are available upon reasonable request to the authors.

## Acknowledgments

This study was supported by grant 5R01DA046224 from the National Institute on Drug Abuse (to Drs Rogers and Agrawal) and grant 5T32MH100019 from the National Institute of Mental Health (to Dr. Constantino-Pettit)

