## Supplemental eTable1, eTable2 for "Associations Between Prenatal Cannabis Exposure and Birth Outcomes: Results from a Prospective Cohort Study"

**RSupplement**

Pg 2: eTable 1. Characteristics of women whose offspring were designated as having prenatal cannabis exposure (PreCE) and those without exposure (nPreCE).

Pg 4: eTable 2. Unadjusted and adjusted (for unadjusted associations at p < 0.05) associations between neonatal outcomes of small (SGA) and large (LGA) for gestational age, Apgar score and NICU admission and prenatal cannabis exposure (PreCE), by frequency (High-PreCE, Low-PreCE), Nicotine presence (PreCE+Nicotine, PreCE-Nicotine) relative to no prenatal cannabis exposure (nPreCE).

**eTable 1: Characteristics of women whose offspring were designated as having prenatal cannabis exposure (PreCE) and those without exposure (nPreCE).**

|  | **PreCE (n= 297)** | **nPreCE (n=151)** | **P value** |
| --- | --- | --- | --- |
| **Maternal age in years (median, [range])** | 25.5 [18.8-40.5] | 27.7 [18.6-40.5] | **0.002** |
| **Advanced maternal age** | 13 (4.4%) | 13 (8.6%) | 0.117 |
| **Teen (18-19yo at T1)** | 24 (8.2%) | 8 (5.3%) | 0.361 |
| **Maternal BMI at intake (kg/m2) (median, [range])** | 27.7 [15.5-67.5] | 30.9 [14.8-79.1] | **0.009** |
| **Education beyond high school** | 81 (29.8%) | 72 (50.7%) | **<0.001** |
| **ADI state decile (median, [range])** | 9.0 [1.0-10.0] | 9.0 [1.0-10.0] | **0.009** |
| **Race** |  |  | **0.022** |
| **American Indian/Alaska Native** | 1 (0.3%) | 1 (0.6%) |  |
| **Asian** | 0 (0%) | 3 (2.0%) |  |
| **Black or African American** | 266 (89.6%) | 122 (80.8%) |  |
| **Hawaiian Native & Pacific Islander** | 0 (0%) | 0 (0%) |  |
| **White** | 28 (9.4%) | 26 (17.2%) |  |
| **Other** | 1 (0.3%) | 1 (0.6%) |  |
| **Ethnicity** |  |  | **0.014** |
| **Hispanic** | 5 (1.7%) | 10 (6.6%) |  |
| **Non-Hispanic** | 291 (98.0%) | 141 (93.4%) |  |
| **Gravidity** **(median, [range])** | 3 [1-7] | 3 [1-7] | 0.729 |
| **Prior live births (median, [range])** | 1 [0-6] | 1 [0-6] | 0.103 |
| **Concurrent medical conditions** |  |  |  |
| **Chronic hypertension** | 65 (22.3%) | 41 (27.9%) | 0.237 |
| **Asthma** | 91 (31.1%) | 46 (30.9%) | 1.000 |
| **Anemia** | 116 (39.6%) | 58 (38.9%) | 0.974 |
| **Psychiatric diagnosis** | 137 (46.8%) | 73 (49.0%) | 0.731 |
| **Prior obstetric outcomes** |  |  |  |
| **Preterm birth** | 44 (22.7%) | 24 (16.0%) | 0.883 |
| **Hypertensive disorders of pregnancy** | 65 (22.3%) | 41 (27.9%) | 0.237 |
| **Gestational diabetes** | 8 (2.8%) | 10 (6.7%) | 0.086 |
| **Cotinine positivity (at any trimester)** | 160 (55.3%) | 18 (12.5%) | **<0.001** |
| **Gestational age at delivery (weeks, [range])** | 38 [24-41] | 38 [26-40] | 0.843 |
| **Attendance at 3 or more prenatal visits** | 284 (97.3%) | 146 (98.0%) | 0.757 |

**eTable 2: Unadjusted and adjusted (for unadjusted associations at p < 0.05) associations between neonatal outcomes of small (SGA) and large (LGA) for gestational age, Apgar score and NICU admission and prenatal cannabis exposure (PreCE), by frequency (High-PreCE, Low-PreCE), Nicotine presence (PreCE+Nicotine, PreCE-Nicotine) relative to no prenatal cannabis exposure (nPreCE).**

|  | **PreCE (n=297)** | **nPreCE (n=151)** | **RR (95% CI), *p*** | ***aRR (95% C.I.)** |
| --- | --- | --- | --- | --- |
| **SGA < 5%ile** | 38 (12.8%) | 6 (4.0%) | 3.22 (1.44-7.33), ***0.005*** | 2.52 (1.06-6.01), ***0.037*** |
| **SGA < 10%ile** | 77 (25.9%) | 19 (12.6%) | 2.06 (1.32-3.28), ***0.002*** | 1.38 (0.83-2.29), *0.208* |
| **LGA** | 7 (2.4%) | 8 (5.3%) | 0.44 (0.17-1.16), *0.175* | NA |
| **5 minute Apgar ≤6** | 9 (3.0%) | 10 (6.6%) | 0.46 (0.20-1.08), *0.129* | NA |
| **NICU admission** | 47 (15.8%) | 23 (15.2%) | 1.05 (0.67-1.66), *0.955* | NA |
|  | **PreCE-High (n=122)** | **PreCE-Low (n=166)** | **RR (95% C.I.)** | ***aRR (95% C.I.)** |
| **SGA < 5ile** | 14 (11.5%) | 22 (13.3%) | 0.9 (0.46-1.60), *0.787* | NA |
| **SGA < 10ile** | 35 (28.7%) | 38 (22.9%) | 1.3 (0.84-1.85), *0.327* | NA |
| **LGA** | 1 (0.0%) | 6 (3.6%) | 0.2 (0.04-1.40), *0.245* | NA |
| **5 min Agpar ≤6** | 2 (1.6%) | 7 (4.2%) | 0.4 (0.09-1.60), *0.310* | NA |
| **NICU admission** | 14 (11.5%) | 29 (17.5%) | 0.7 (0.36-1.16), *0.206* | NA |
|  | **PreCE-High (n=122)** | **nPreCE (n=151)** | **RR (95% C.I.)** | ***aRR (95% C.I.)** |
| **SGA < 5ile** | 14 (11.5%) | 6 (4.0%) | **2.9 (1.18-7.10), *0.033*** | **2.9 (1.07-8.06), *0.029*** |
| **SGA < 10ile** | 35 (28.7%) | 19 (12.6%) | **2.3 (1.39-3.77), *0.002*** | 1.6 (0.89-2.87), *0.095* |
| **LGA** | 1 (0.0%) | 8 (5.3%) | **0.2 (0.03-0.93), *0.045*** | 0.1 (0.00-2.54), *0.210* |
| **5 min Agpar ≤6** | 2 (1.6%) | 10 (6.6%) | **0.2 (0.06-0.98), *0.071*** | 0.2 (0.03, 1.35), *0.087* |
| **NICU admission** | 14 (11.5%) | 23 (15.2%) | 0.8 (0.41-1.39), *0.478* | NA |
|  | **PreCE-Low (n=166)** | **nPreCE (n=151)** | **RR (95% C.I.)** | ***aRR (95% C.I.)** |
| **SGA < 5ile** | 22 (13.2%) | 6 (4.0%) | **3.3 (1.44-7.84), *0.007*** | 2.3 (0.93-5.79), *0.071* |
| **SGA < 10ile** | 38 (22.9%) | 19 (12.6%) | **1.8 (1.11-3.01), *0.025*** | 1.3 (0.72-2.27), *0.392* |
| **LGA** | 6 (3.6%) | 8 (5.3%) | 0.7 (0.25-1.84), *0.649* | NA |
| **5 min Agpar ≤6** | 7 (4.2%) | 10 (6.6%) | 0.6 (0.26-1.59), *0.498* | NA |
| **NICU admission** | 29 (17.5%) | 23 (15.2%) | 1.2 (0.71-1.91), *0.669* | NA |
|  | **PreCE+Nicotine (n=157)** | **PreCE-Nicotine (n=133)** | **RR (95% C.I.)** | ***aRR (95% C.I.)** |
| **SGA < 5ile** | 27 (17.2%) | 11 (8.3%) | **2.1 (1.09-4.01), *0.038*** | **2.2 (1.11-4.26), *0.016*** |
| **SGA < 10ile** | 52 (33.1%) | 23 (17.3%) | **1.9 (1.25-2.96), *0.003*** | **2.0 (1.24-3.06), *0.002*** |
| **LGA** | 1 (0.0%) | 6 (4.5%) | **0.1 (0.02-0.88), *0.050*** | 0.2 (0.02-1.37), *0.080* |
| **5 min Agpar ≤6** | 4 (2.5%) | 5 (3.8%) | 0.7 (0.21-2.34), *0.738* | NA |
| **NICU admission** | 23 (14.6%) | 22 (16.5%) | 0.9 (0.53-1.52), *0.805* | NA |
|  | **PreCE+Nicotine (n=157)** | **nPreCE (n=123)^&^** | **RR (95% C.I.)** | ***aRR (95% C.I.)** |
| **SGA < 5ile** | 27 (17.0%) | 6 (4.8%) | **3.6 (1.57-8.23), *0.003*** | **3.5 (1.41-8.52), *0.027*** |
| **SGA < 10ile** | 54 (34.0%) | 16 (12.7%) | **2.7 (1.64-4.45), *<0.001*** | **2.3 (1.35-3.94), *0.007*** |
| **LGA** | 1 (0.1%) | 6 (4.8%) | **0.1 (0.02-0.82), *0.046*** | 0.2 (0.02-1.28), *0.171* |
| **5 min Agpar ≤6** | 4 (2.5%) | 8 (6.3%) | 0.4 (0.13-1.24), *0.142* | NA |
| **NICU admission** | 23 (14.5%) | 18 (14.3%) | 1.0 (0.58-1.80), *1.000* | NA |
|  | **PreCE-Nicotine (n=133)** | **nPreCE (n=123)^&^** | **RR (95% C.I.)** | ***aRR (95% C.I.)** |
| **SGA < 5ile** | 11 (8.0%) | 6 (4.7%) | 1.7 (0.66-4.26), *0.402* | NA |
| **SGA < 10ile** | 23 (16.7%) | 16 (12.7%) | 1.3 (0.74-2.36), *0.436* | NA |
| **LGA** | 6 (4.3%) | 6 (4.7%) | 0.9 (0.32-2.63), *1.000* | NA |
| **5 min Apgar ≤6** | 5 (3.6%) | 8 (6.3%) | 0.6 (0.20-1.61), *0.397* | NA |
| **NICU admission** | 24 (17.4%) | 18 (14.3%) | 1.2 (0.70-2.12), *0.951* | NA |
| *Abbreviations: aRR, adjusted relative risk ; CI, confidence interval; N/A, not applicable; LGA, large for gestational age; NICU, neonatal intensive care unit; PreCE+Nicotine, prenatal cannabis use with cotinine positivity; PreCE-Nicotine, prenatal cannabis use with negative cotinine; RR, relative risk; SGA, small for gestational age*  **aRR for neonatal outcomes were adjusted for: maternal age, maternal BMI, cotinine positivity, ADI, infant gestational age at birth; see Figure 1 for adjusted linear models with continuous birthweight outcome*  ***^&^****N=18 women who did not report cannabis use or test urine positive for THC-COOH (i.e., nPreCE) had a urine positive for cotinine and were excluded from these comparisons.* | | | | |
